# Efficacy and safety of stem cell therapy for diabetic kidney disease: a systematic review and meta-analysis

**DOI:** 10.1101/2024.11.07.24316903

**Authors:** Hongyu Du, Chen Xie, Yiqin Yuan, Yun Luo, Jinguo Cao, Zhihai Li, Jiayi Yuan, Wei Li

**Affiliations:** Gannan Medical University, Ganzhou, Jiangxi 341000, China

**Keywords:** stem cells, diabetic nephropathy, clinical trial, meta-analysis, systematic review, randomized controlled trials

## Abstract

**Background/Objectives:** Animal studies have demonstrated the ability of stem cell therapy (SCT) to treat diabetic kidney disease (DKD). However, the safety and efficacy of SCT in patients with DKD remain unclear. This systematic review and meta-analysis aimed to investigate the safety and efficacy of SCT in patients with DKD.

**Methods:** A comprehensive and systematic literature search was conducted using PubMed, EMBASE, Cochrane Library, and Web of Science to identify articles on SCT for DKD published up to March 2024. RevMan V.5.4 software was used for statistical analysis.

**Results:** We identified four studies that included 90 participants, 53 (58%) of whom underwent SCT. SCT improved estimated glomerular filtration rate (eGFR) (mean difference [MD] = 0.41, 95% confidence interval [CI]: 0.08–0.74; p < 0.05), serum creatinine (SCr) reduction (standardized MD = -0.65, 95%CI: -1.19 to -0.1, p < 0.05), and microalbuminuria (MAU) (MD = -32.10, 95% CI: -55.26–8.94; p < 0.05) compared to the control group, but did not improve urine microalbumin/creatinine ratio (UACR) (MD = -63.36, 95% CI: 194.52–67.79, p = 0.56) or blood sugar (MD = 0.49, 95% CI: 4.16–2.01, p = 0.49). In addition, the incidence of adverse events was significantly high in both groups (risk ratio = 0.93; 95% CI: 0.74–1.17; p = 0.54); there was no significant difference regarding I_2_ = 0%).

**Conclusions:** SCT can safely and effectively improve eGFR and SCr levels by lowering the MAU but cannot improve UACR and blood sugar levels.

---

Diabetes mellitus (DM) is a chronic disease with a high incidence rate [1] and remains a major global public health challenge [2–6]. According to statistics, 25–40% of patients with diabetes develop secondary diabetic nephropathy (DN) and, eventually, end-stage kidney disease (ESKD) [7], an irreversible phase in which the kidneys completely lose the ability to filter waste products and excess fluids. Consequently, patients rely on dialysis or kidney transplantation to sustain life, along with a high risk of cardiovascular disease and death [8].

Stem cell therapy (SCT) is a promising biotechnology technique with wide applications and has made remarkable advances in clinical settings [9]. The stem cells that have been used in preclinical and clinical studies include umbilical cord blood mesenchymal stem cells (MSCs) [10], umbilical cord MSCs [11], placental MSCs [12], adipose MSCs [13], and bone marrow mesenchymal stromal cells (BMSCs) [14]. Among them, adipose MSCs are the most widely used. Animal experiments have shown that SCT can effectively treat diabetic kidney disease (DKD) [15,16]. However, the safety and efficacy of SCT in patients with DKD remain unknown, and only a few randomized controlled trials (RCTs) with small sample sizes have explored its role in the treatment of patients with DKD. Norberto et al. [17] recently conducted a phase 1b/2a multicenter RCT to evaluate the safety, tolerability, and treatment efficacy of adult allogeneic bone marrow stromal stem cell transplantation in patients with moderate-to-severe DKD. The authors found that 18 weeks of SCT resulted in significant improvement of estimated glomerular filtration rate (eGFR) but did not affect the urine microalbumin/creatinine ratio (UACR). However, Gaipov et al. [18] found that SCT significantly reduced microalbuminuria (MAU) without affecting eGFR or serum creatinine (SCr) levels. Therefore, this systematic review and meta-analysis of RCTs aimed to explore the safety and efficacy of SCT in patients with DKD to provide deeper insights into the translation of SCT from clinical trials to the clinical application stages.

## 2. Methods

### 2.1. Protocol and Registration

This systematic review and meta-analysis has been registered in PROSPERO (CRD42021282869) and was performed according to the Preferred Reporting Items for Systematic Reviews and Meta-Analyses (PRISMA) guidelines [19].

### 2.2. Search Strategy

As of March 4, 2024, two authors (H-Y.D. and C.X.) comprehensively retrieved clinical trial data relating to renal-related outcome measures and adverse events in adults with DKD using PubMed, Cochrane Library, Web of Science, and Embase to assess SCT efficacy. The main search terms were “stem cells,” “Diabetic Nephropathies,” “Diabetic Kidney Diseases,” and related keywords. Full details of the retrieval strategy for all the databases can be found in the Supplementary Materials.

### 2.3. Inclusion Criteria

1. Population: age ≥ 18 years old; established diagnosis of type 1 or type 2 DM with DKD; eGFR < 60 mL/min/1.73 m_2_ for three consecutive months or MAU (albumin of 30–300 mg in a 24-h urine collection). DN was defined as either micro- or macroalbuminuria (albumin > 300 mg/24-h) according to the 2007 Kidney Disease Outcomes Quality Initiative Clinical Practice Guidelines and Clinical Practice Recommendations for Diabetes and Chronic Kidney Disease [20]. All studies of patients with DKD that reported at least one of the following results were considered for inclusion: UACR, cystatin C, SCr, eGFR, markers of tubular injury, adverse event rate, and mortality.
2. Intervention: stem cell drug products, regardless of source, type, dose, duration, or route of administration.
3. Comparison intervention: placebo. Trials with multiple interventions (e.g., co-administered autologous bone marrow-derived mononuclear cells and umbilical cord-MSCs) were eligible if the study groups differed only in their use of SCs.
4. Outcome(s): the primary outcome was eGFR; the secondary outcomes were SCr, MAU, UACR, and incidence of adverse events; other relevant outcome measures included metabolic parameters: hemoglobin A1c, triglycerides, and glucose.
5. Language: All articles published in English.

### 2.4. Exclusion Criteria

The exclusion criteria were as follows: (1) animal experiments; (2) kidney disease secondary to other diseases; (3) full-text content not available; and (4) missing or duplicated experimental data.

### 2.5. Study Selection

After removing duplicate studies, two authors (H-Y.D. and C.X.) independently screened all titles and abstracts for potential relevance and acquired the full text of the relevant content. Disagreements were resolved by consensus or by consulting a third author (Y.Y.).

### 2.6. Data Extraction and Literature Quality Evaluation

#### 2.6.1. Date Collection

Two authors (H-Y.D. and C.X.) summarized the primary data from the included trials, including the first author and year of publication. If the data were not reported or missing, the corresponding author was emailed. If the authors did not respond, data were obtained from the charts or formulas. Disagreements were resolved by consulting a third author (Y.Y.).

#### 2.6.2. Assessment of Risk of Bias and Quality of Evidence

The quality of each study included in the analysis was assessed using the Cochrane Risk of Bias Assessment Tool (RevMan 5.40 There were seven items in the bias risk table: (1) random sequence generation (selection bias); (2) allocation concealment (selection bias); (3) blinding of participants and personnel (performance bias); (4) blinding of outcome assessment (detection bias); (5) incomplete outcome data (attrition bias); (6) selective reporting (reporting bias); and (7) other bias. Each item was classified as low risk, high risk (not fulfilling the criteria), or unclear (specific details or descriptions were not reported) [21]. Furthermore, the presence of publication bias was estimated using a funnel plot.

### 2.7. Data Analysis

Review Manager (5.40; Cochrane Collaboration) software was used for statistical analysis. Between-study heterogeneity was assessed using the Higgins I_2_-test. Meaningful heterogeneity was determined at 50% of the I_2_ values. Due to significance, a random-effects model was used for the meta-analysis over a fixed-effects model. For dichotomous variable data such as mortality, the risk ratio (RR) and 95% confidence interval (95% CI) were used as the combined effect size estimates. For continuous variables, such as eGFR and SCr, standardized mean difference (SMD) or weighted mean differences and their 95% CI were used as the combined effect size estimates.

G1, experimental group; CG, control group; ADN, advanced diabetic nephropathy; EDN, early diabetic nephropathy; MPC, mesenchymal precursor cells; ABM-MNCs, autologous bone marrow-derived mononuclear cells; UC-MSCs, umbilical cord mesenchymal stem cells; I.V: intravenous injection, I.A: arterial injection, TEAEs: treatment-emergent adverse events

AE, adverse event; M, months; Y, years; mGFR, measured glomerular filtration rate; eGFR, estimated glomerular filtration rate; MDRD, Modification of Diet in Renal Disease; CKD-EPI, Chronic Kidney Disease Epidemiology Collaboration; UACR, urine albumin/creatinine ratio; HbA1c, hemoglobin A1c; LDL, low-density lipoprotein; BP, blood pressure; HLA, ; DPN, ; DN, diabetic nephropathy; DRP, ; SCr, serum creatinine; IL, interleukin; TNF, tumor necrosis factor; TGF, transforming growth factor; FGF23, NGAL, ; ACE,

**Table 1.** Study characteristics.

| First author, year | Injection Dose | Duration | Primary Outcomes | Secondary Outcomes |
| --- | --- | --- | --- | --- |
| Norbert o, 2023 | 80×10<sup>6</sup> cells | 18M | Safety: the number and severity of prespecified cell infusion-associated events and the overall number and frequency of AEs and unexpected severe AEs | mGFR, eGFR (MDRD, CKD-EPI), UACR, fasting blood glucose, HbA1c, total cholesterol, LDL cholesterol, triglycerides, BP, anti-HLA antibody development, proportion of total number of circulating lymphocyte (T cells, B cells, and NK cells), myeloid cell (monocytes and dendritic cells) subsets and plasma serum immunoassay-derived concentrations of biomarkers of inflammation |
| Zhixian Wu, 2021 | 1.10 ± 0.22×10<sup>6</sup> MSCs/kg<br>0.61 ± 0.26×10<sup>10</sup> aBM-MNCs/kg | 8Y | The incidence of chronic complications, including DPN, DN, DRP. | Safety, HbA1c, exogenous insulin requirement (daily dose), fasting blood glucose, fasting C peptide, microalbumin, SCr, eGFR (MDRD) |
| Packham, 2016 | -<br>150×10<sup>6</sup> cells<br>300×10<sup>6</sup> cells | 60M | Safety, eGFR (MDRD, CKD-EPI), mGFR | Serum creatinine, Creatinine clearance, Albumin-creatinine ratio, Protein-creatinine ratio, Cystatin-C, HbA1c or BP, IL-6, TNF-α, adiponectin, TGF-β, uric acid, FGF23 |
| Gaipov, 2018 | 140×10<sup>6</sup> cells |  | NGAL, Urinary type-IV collagen, microalbuminuria, eGFR (CKD-EPI) | Fasting C-peptide, fasting serum insulin, HbA1C, glucose fasting, glucose postprandial, insulin-replacement, insulin short-acting, insulin long-acting, β-blockers, ACE-inhibitors, Ca-channel blockers |

AE, adverse event; M, months; Y, years; mGFR, measured glomerular filtration rate; eGFR, estimated glomerular filtration rate; MDRD, Modification of Diet in Renal Disease; CKD-EPI, Chronic Kidney Disease Epidemiology Collaboration; UACR, urine albumin/creatinine ratio; HbA1c, hemoglobin A1c; LDL, low-density lipoprotein; BP, blood pressure; HLA, ; DPN, ; DN, diabetic nephropathy; DRP, ; SCr, serum creatinine; IL, interleukin; TNF, tumor necrosis factor; TGF, transforming growth factor; FGF23, NGAL, ; ACE,

## 3. Results

### 3.1. Eligible Studies

The PRISMA flow diagram is shown in Figure 1. A systematic electronic literature search initially identified 3,528 studies. After applying the exclusion criteria, four trials [17,18,22,23] were included in the meta-analysis.

**Figure 1.**
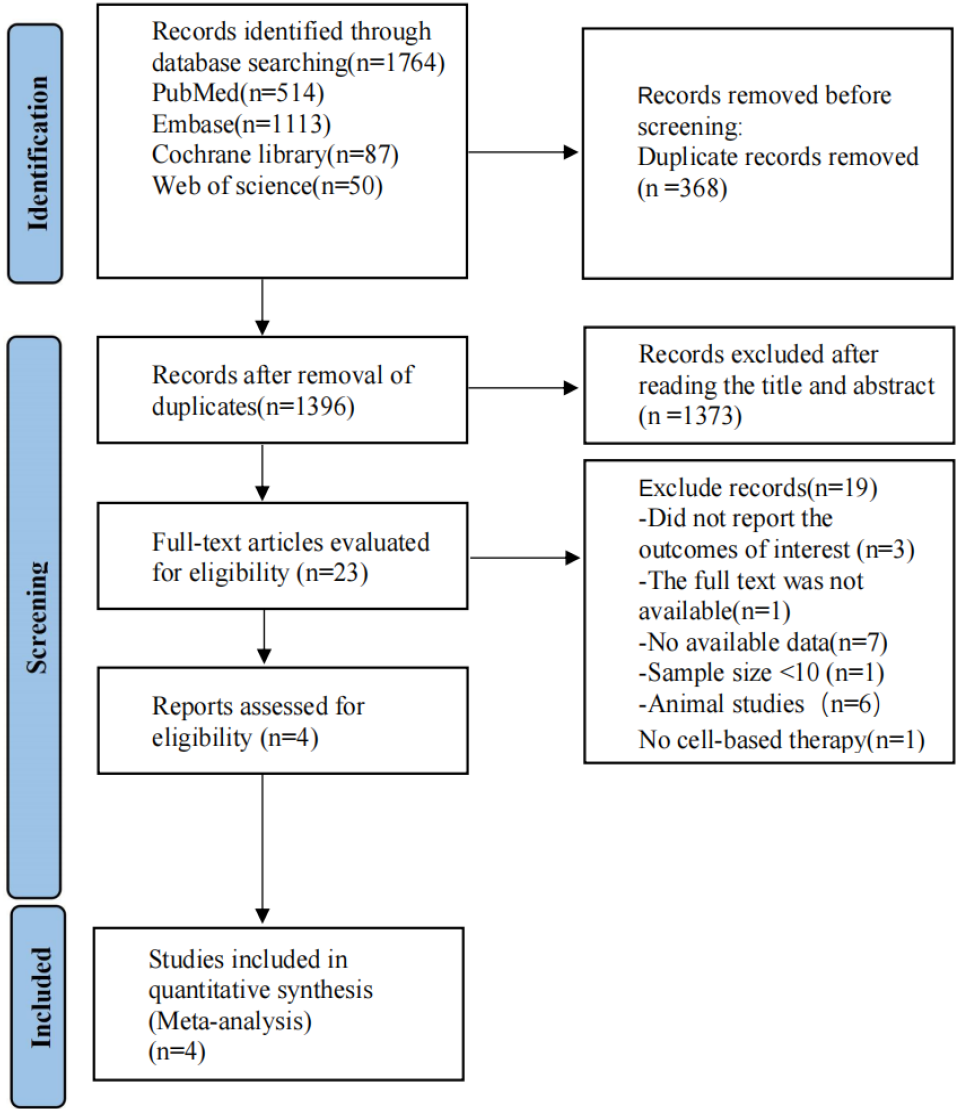
PRISMA flow diagram of the study process. PRISMA, Preferred Reporting Items for Systematic review and Meta-analysis

### 3.2 Study Characteristics

Baseline data and interventions are presented in Table 1,2. The included studies were published between 2016 and 2023, with four articles and 90 participants. A total of 53 (58%) patients underwent SCT. Although SCT was applied in all included studies, the source, dose, frequency, and mode of injection varied. All four studies included used bone marrow as a source of stem cells, and one study used umbilical cord MSCs. Allogeneic administration was employed in two studies, and autologous administration was employed in two studies.

**Table 1.**
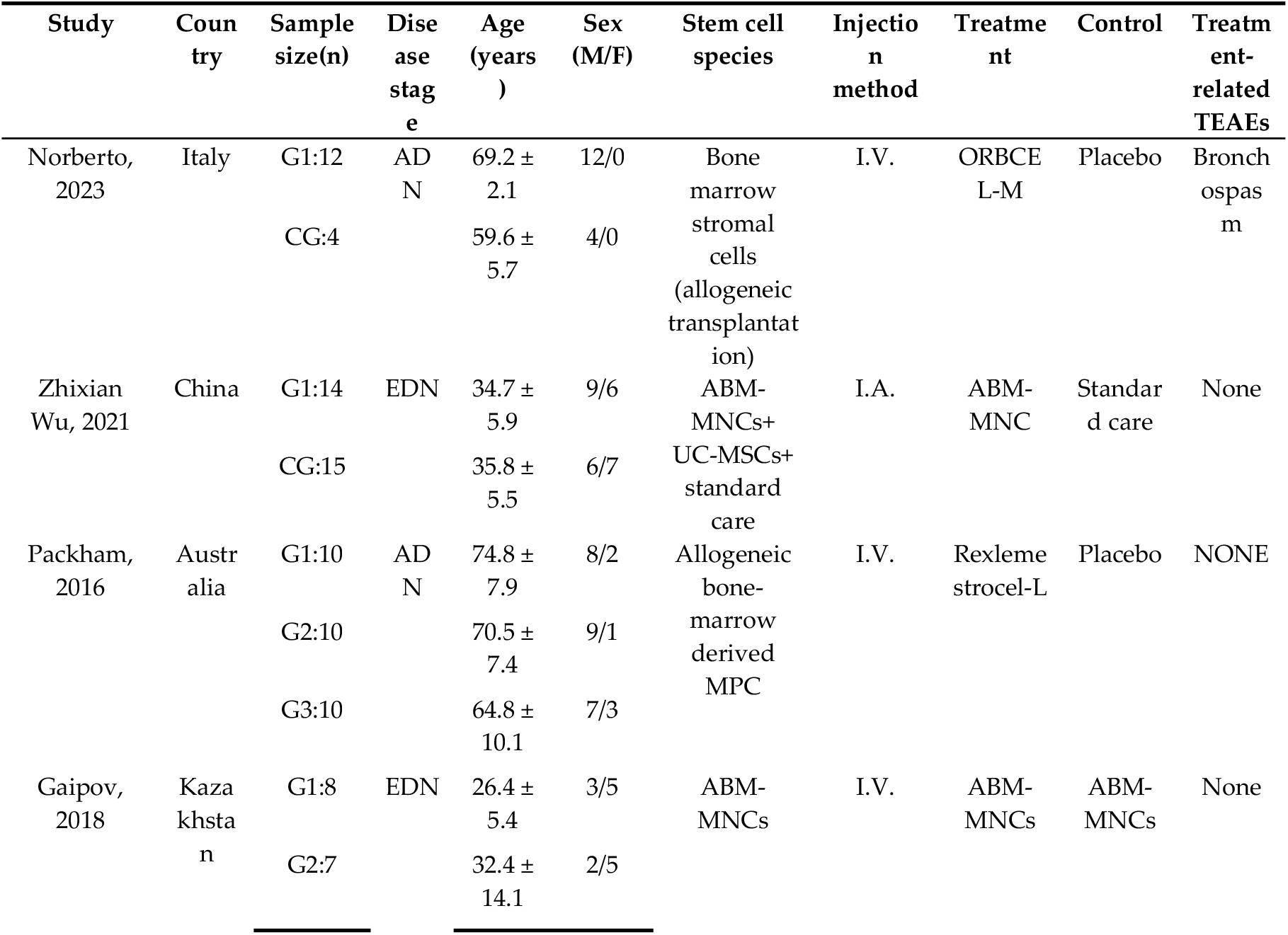
Patient Characteristics

### 3.3. Quality Assessment of the Articles

Figures 2 and 3 summarize the risk of bias in the included studies. The four studies had different study designs; three studies were RCTs [17,18,22], and one was a prospective, open-label study [23]. Furthermore, quality assessment of these studies revealed that three studies had a low risk of bias [17,18,22], and one study had an unclear to high risk of bias, as its investigators did not apply the blinding procedure rationally [23]. Overall, the included RCTs had a low risk of bias.

**Figure 2.**
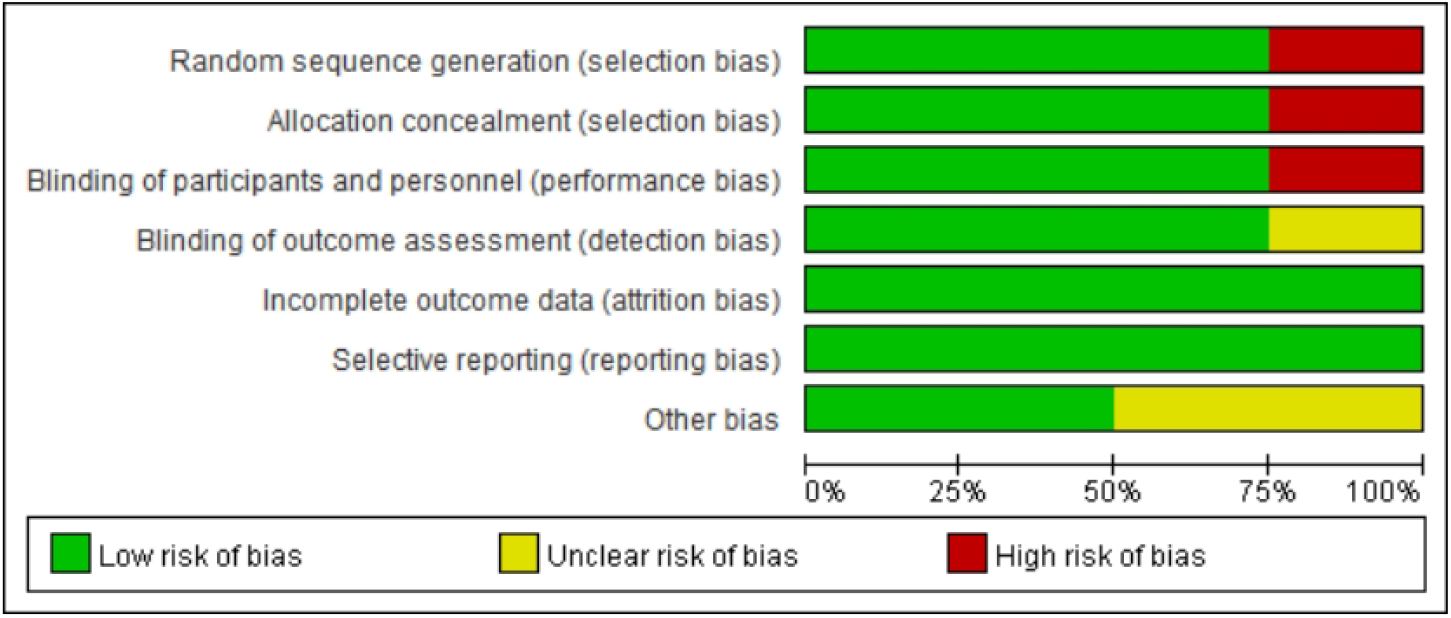
Risk of bias graph: judgments of each risk of bias item, presented as a percentage.

**Figure 3.**
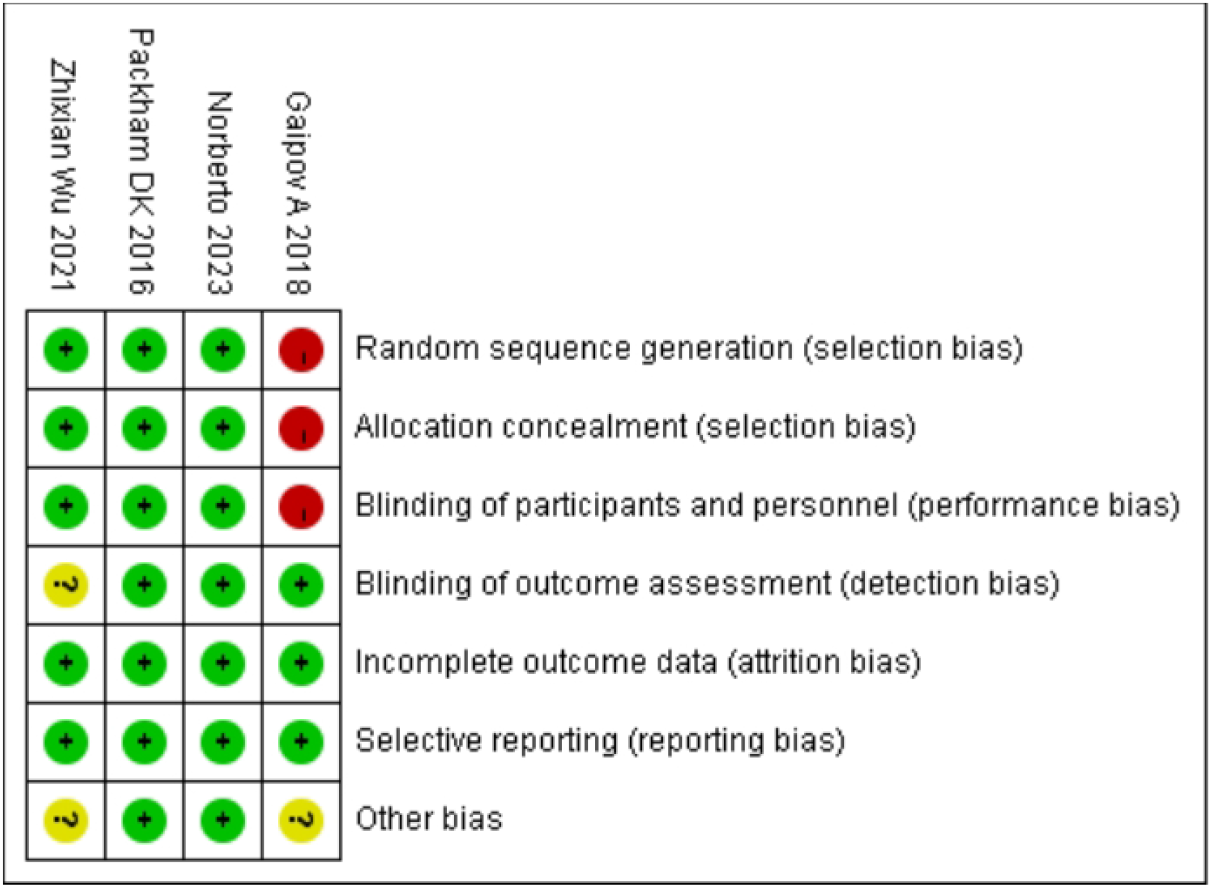
Risk of bias summary: judgments of each risk of bias item for each included study

### 3.4. Outcome

#### 3.4.1. Effect of SCT on eGFR

The eGFR is an important indicator of renal function. The Modification of Diet in Renal Disease and Chronic Kidney Disease Epidemiology Collaboration formulas are commonly used to estimate GFR. Four studies [17, 22, 23] showed that SCT significantly improved eGFR levels (Z = 3.56; p = 0.02). Analysis of forest plot data (Fig. 4) showed significant improvement with SCT as the intervention, compared with the outcome in the control group (MD = 0.41, 95% CI: 0.08–0.74; p < 0.05).

**Figure 4.**
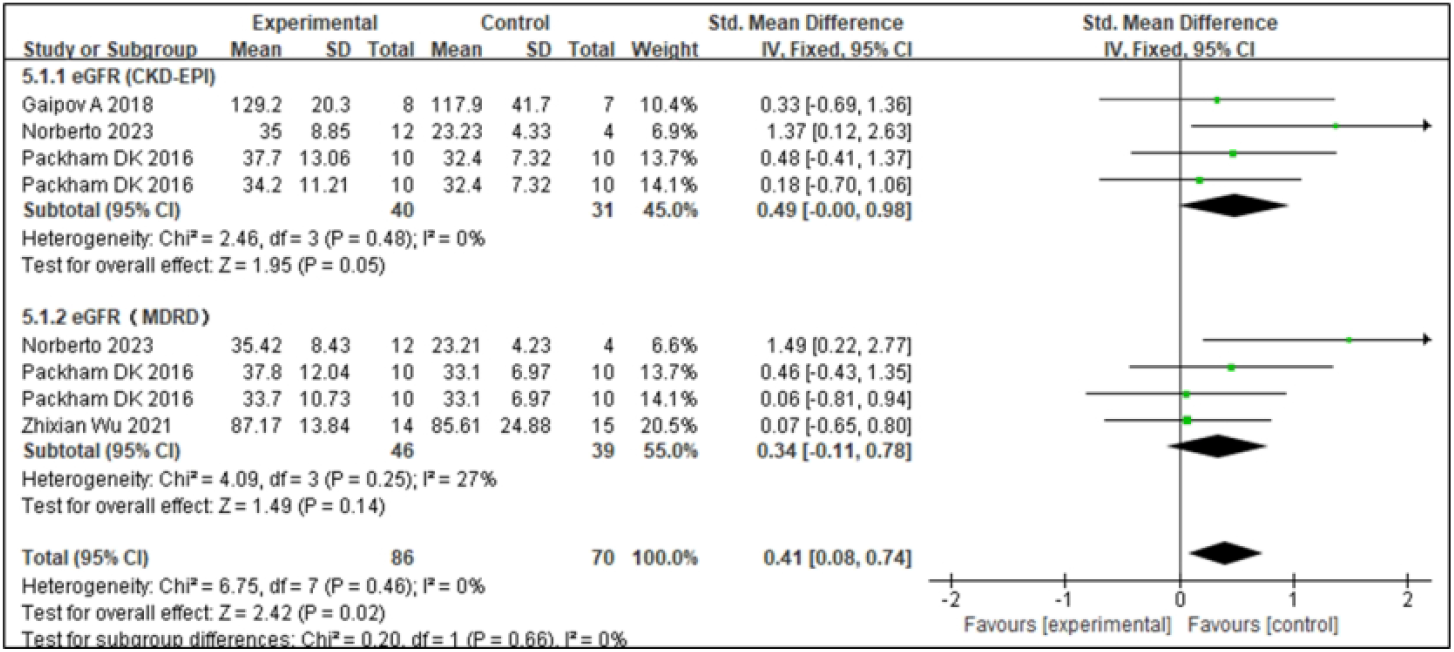
Subgroup analysis for estimated glomerular filtration rate (eGFR)

#### 3.4.2. Effect of SCT on SCr

SCr levels can be used to monitor DKD. In the early stages of DKD, SCr may remain within the normal range, but its levels gradually increase with disease progression; therefore, monitoring SCr levels is important for the early diagnosis and disease monitoring of DKD. Three studies [17,18,23] reported SCr levels, and the associated I_2_ value was 0%. Therefore, we used a fixed-effects model in this study. The results from the forest plot analysis (Fig. 5) showed that treatment with SCT was associated with significant changes in SCr levels (Z = 2.34; p = 0.02), and the trial group with stem cell injection as the intervention showed significantly reduced SCr levels in patients with diabetes (SMD = -0.65, 95% CI= -1.19 to - 0.1, p < 0.05).

**Figure 5.**
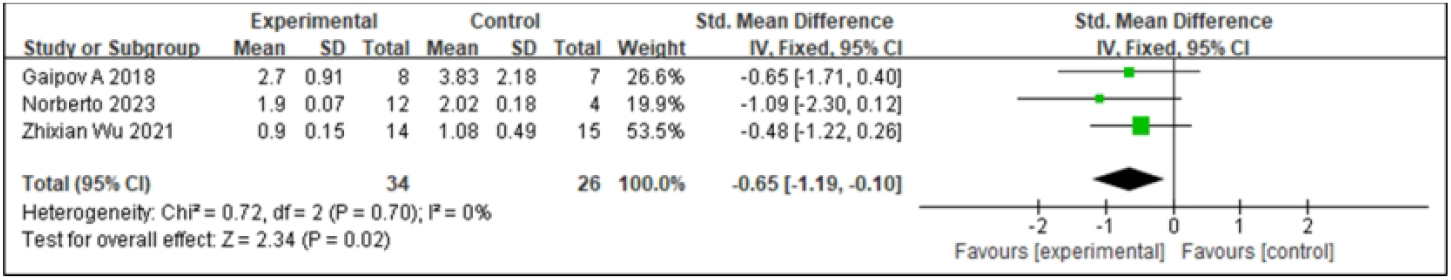
Forest plot for serum creatinine (SCr)

#### 3.4.2. Effect of SCT on MAU

MAU is an early hallmark of DKD. Persistent MAU was significantly positively associated with the risk of developing clinical proteinuria in patients with diabetes, indicating that MAU is important for preventing DKD development. A comprehensive analysis of MAU was conducted in two studies [18,23], presented in Figure 6A. SCT was associated with significant changes in MAU levels (Z = 2.72; p = 0.007), with low inter-study heterogeneity and an I_2_ value of 41, suggesting high agreement between the findings. Analysis of the forest plot data showed that MAU levels in the SCT group were significantly lower than those in the control group (MD = -32.10,95%CI: -55.26 to -8.94; p < 0.05).

**Figure 6.**
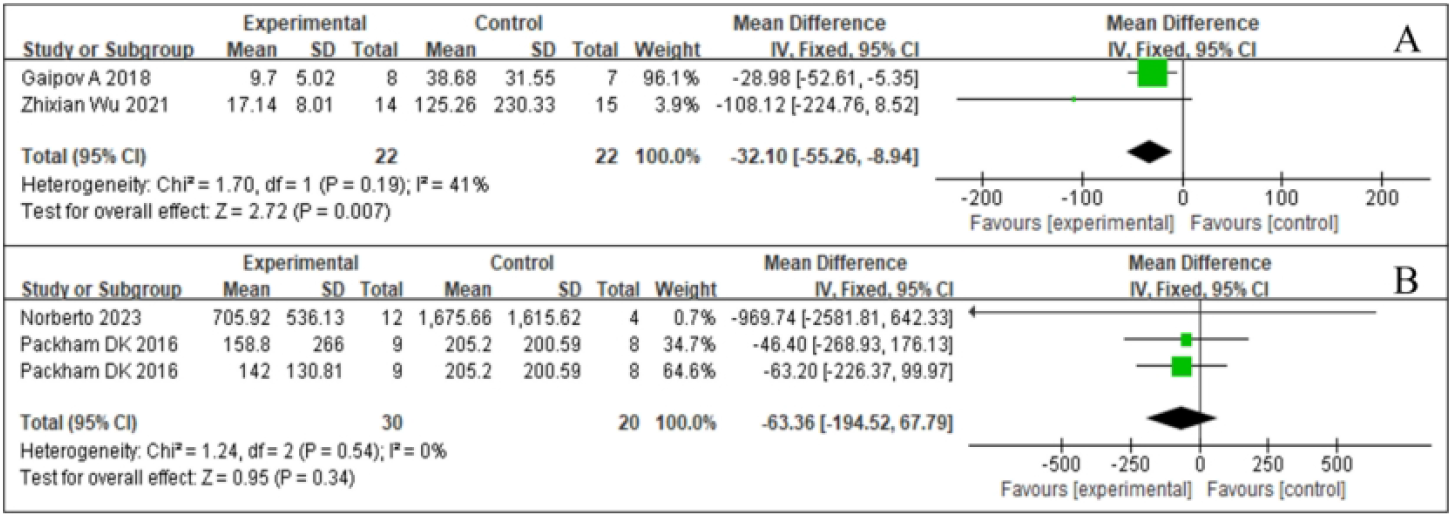
Forest plot for urine markers: A: microalbuminuria (MAU); B: urine albumin/creatinine ratio (UACR).

#### 3.4.5. Effect of SCT on UACR

UACR is an indicator for urinary protein excretion and is a key parameter in the early screening of DKD. Elevated UACR predicts the presence of kidney injury, especially in patients with diabetes. Integrating the available data [18,23], we visually demonstrated the results of the UACR study (Fig. 6B). After statistical analysis, the effect of SCT in reducing UACR did not meet the requirements of statistical significance (Z = 1.51, p = 0.13). In addition, inter-study heterogeneity was low (I² = 0%, p = 0.54). No significant difference in the UACR was found between the test and control groups (SMD = 0.45, 95%CI: -1.05 to 0.14, p = 0.13).

#### 3.4.6. Adverse Events

Regarding the safety of injected stem cells, we performed a meta-analysis of the studies [17,22,23] with respect to the SCT-induced adverse effects and observed no significant difference in any adverse event or serious adverse event (RR = 0.93; 95% CI: 0.74–1.17; p = 0.54; I_2_ = 0%) (Fig. 7).

**Figure 7.**
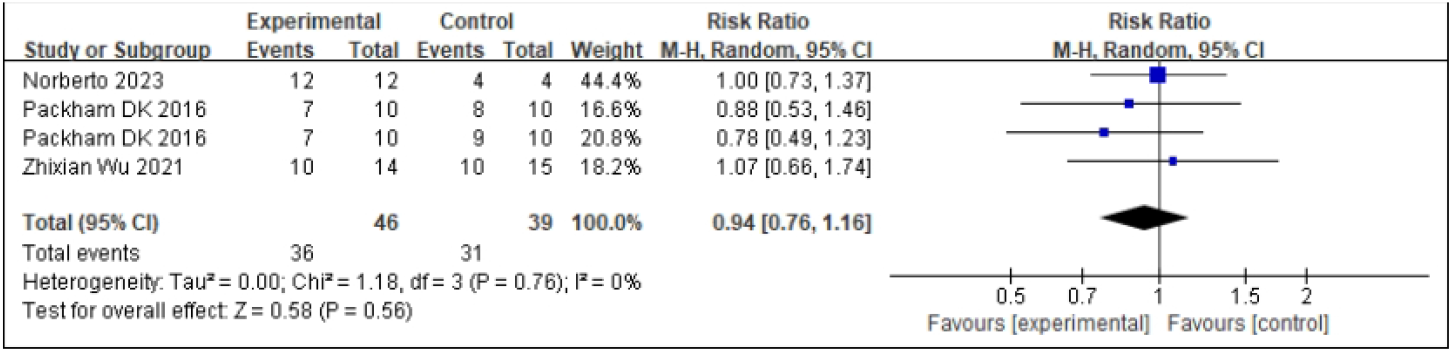
Forest plot of the comparison of the adverse event incidence between the stem cell therapy (SCT) and control groups.

## 4. Discussion

A previous systematic review demonstrated the significant effect of SCT on chronic kidney disease in animal models by showing that it can help reduce the incidence of DKD. This treatment effectively improved kidney function while reducing the release of kidney injury markers, renal fibrosis, and inflammatory mediators, as well as high glucose levels, MAU, eGFR, and SCr levels [15,16,24]. Previous studies have largely been based on these models; however, the efficacy and safety of SCT for DKD remain nebulous owing to the lack of long-term clinical trial data. In particular, the types of stem cells, their sources, and the selection of dosages are controversial among different studies. In this study, we included four RCTs and found that SCT safely and effectively improved eGFR and SCr levels and reduced MAU in patients with DKD. However, SCT did not improve UACR or blood sugar levels (Supplemental Figure 1). Additionally, there was no significant difference in the incidence of adverse events between the two groups.

The efficacy of SCT through various cell delivery pathways and in various cell types remains controversial. Intravenous delivery of MSCs, currently the most widely studied cell type for DKD and related kidney diseases, is restricted by the lungs and spleen, which results in a low number of cells reaching the kidney that may not be sufficiently active [25]. Following the intravenous infusion, most MSCs remain in the lungs in the short term, with 50–60% of MSCs remaining in the lungs at 1 h post-injection, decreasing to 30% after 3 h, and maintaining stable levels at 96 h [26]. Subsequently, the MSCs are gradually cleared from the lungs and accumulate in the liver and spleen. This phenomenon is known as the “lung first-pass effect” [27]. Due to their large size, MSCs are easily trapped in the lung capillaries. Therefore, different infusion routes or preconditioning methods may increase the number and activity of MSCs reaching the kidney.

Other types of cells may have better results in improving kidney outcomes. For example, UC/AF cells reduce SCr, fibrosis, and inflammation similar to MSCs and to a greater extent than by non-MSCs [28]. Compared to MSCs, UC/AF cells also reduced proteinuria to a greater extent. Arterial injection can avoid pulmonary entrapment in the first cycle and improve the targeting efficiency. Researchers have examined the efficacy of various cell delivery pathways in animal models of chronic kidney disease. In a meta-analysis, the caudal vein (70% of studies, 28 animals) was the most effective in reducing renal function outcomes; however, in one study, renal artery delivery was more effective in reducing anti-fibrotic factors than previously reported. Rashed et al. [29] and Han et al. [30] have shown that melatonin(MT)preconditioning can improve the proliferative antioxidant capacity and angiogenesis capacity of BMSCs and enhance their therapeutic effect on DN by promoting the recovery of neurotrophic effects and myelination. These methods may increase the accumulation of MSCs in the kidneys, thereby enhancing their therapeutic effect. In our meta-analysis, all cells were MSCs, and only one of the included studies [23] used arterial injections. However, there was no significant difference in SCr or eGFR levels between the SCT and control groups, unlike in MAU levels. Future studies may provide a clear answer regarding the superior cell injection pathways and cell tissue sources in DKD therapy.

The two most effective biomarkers for assessing kidney health are eGFR and albuminuria (or proteinuria) [31]. eGFR is the gold standard for accurately measuring overall kidney function [32]. In addition, estimates of eGFR are based on serological biomarkers of renal filtration, most commonly SCr [33]. In existing animal models and clinical trials, SCT is associated with improvements in renal function, such as stabilization or enhancement of GFR and reduction of proteinuria. Lin et al., [15] in a meta-analysis, found that SCT has a potential renoprotective effect, significantly reducing SCr and blood urea nitrogen levels and mitigating renal impairment. The meta-analysis by Papazova et al. [16] showed that SCT could reduce the occurrence and progression of chronic kidney disease, especially through the improvement of urinary protein, SCr, and eGFR levels. The results of the meta-analysis in this study are consistent with these findings, showing that SCT significantly improved the degree of disease activity, albuminuria, SCr, and eGFR levels in DKD. However, the GFR level at which individuals benefit the most from SCT remains undetermined, and this “treatment window” has been explored in clinical nephrology trials, including the angiotensin-receptor blocker irbesartan in DN [34] and fish oil in IgA nephropathy [35].

In addition, we found differences in the efficacy of cell therapies at the molecular level as well as changes in blood glucose levels between different species, which may be related to the tightly controlled conditions and detailed evaluation of animal trials. Clinical trials must consider more practical application factors, such as individual differences and concomitant diseases. For example, Ezquer et al. [36] administered pluripotent mesenchymal stromal cells to mice with DM to study the preventive effect of SCT on chronic kidney disease secondary to DM; this led to the regeneration of the pancreas and kidneys by reversing high blood sugar levels and reducing proteinuria. In another study, Ezquer et al. observed a reduction in proteinuria despite hyperglycemia and hypoinsulinemia following transplantation of autologous bone marrow mesenchymal stem cells(AB-MSCs), highlighting the direct renoprotective function of stem cells [37]. The opposite results were obtained by Zhou et al. [38]. DN was induced in Sprague-Dawley rats using intrabitoneal injection of streptozotocin, and, after MSC transplantation, the blood glucose level showed improvements but proteinuria did not improve. Wang et al. [39] investigated direct renal regeneration in experimental rat models with type 1 DN, where intra-arterial administration of BMSCs prevented the development of proteinuria and podocyte damage or loss but did not improve blood glucose levels. In the current meta-analysis, SCT treatment was significantly effective in reducing albuminuria but not in improving glycemic control in patients with DKD. This result should be interpreted with caution as it is based on pooled data from a small number of studies.

MSC infusion reduces the production of profibrotic markers and inflammatory factors, as demonstrated by decreased levels of interleukin (IL)-6 and tumor necrosis factor-α (TNF-α) and increased levels of the anti-inflammatory cytokines IL-4 and IL-10 [40,41]. Li et al. [26] determined the levels of validated cytokines in serum samples of DN rats using Milliplex rat cytokine kit and suggested that MSC treatment significantly reduced the expression of IL-1α, IL-1β, IL-6, and interferon-γ. After lipopolysaccharide stimulation of macrophages, the expression of proinflammatory cytokines such as IL-6, IL-1β, TNF-α, and monocyte chemoattractant protein-1 increased. SCT can also serve as treatment for other kidney diseases. Chang et al. [42] evaluated the role of MSCs in anti-Thy1.1-induced glomerulonephritis rat models and found that the intrarenal transplantation of hypoxia-preconditioned MSCs reduced glomerular apoptosis, autophagy, and inflammation. Song et al., [43] in adriamycin (ADR) nephropathy rats, showed that MSCs reduced oxidative stress and inflammation by inhibiting nuclear factor-kappa B and improved glomerular sclerosis and interstitial fibrosis, alleviating ADR nephropathy. In the clinical trials included in this study, SCT did not have a prominent anti-inflammatory effect; Norberto et al.’s trial [17] showed an increasing trend in the serum inflammatory biomarkers such as soluble TNF receptor 1, neutrophil gelatinase-associated lipocalin, and vascular cell adhesion molecule 1 during the 18-month follow-up period, with no difference between groups. A multicenter RCT study by Packham et al. [22] showed no significant change in TNF-α levels. Owing to the differences in the anti-inflammatory effects of MSCs observed in animal models and clinical trials, the inflammatory markers selected in different studies may differ, and the measurement methods may affect the interpretation of the results. For example, some studies may use more sensitive biomarkers or more precise measurement techniques that more accurately reflect changes in the inflammatory status. In animal studies, the route of administration of MSCs (intravenous injection and intrarenal transplantation) and dosage may differ from those in clinical trials. In humans, higher doses of MSCs or specific routes of administration may be required to achieve anti-inflammatory effects similar to those observed in animals.

Exploring the potential mechanisms underlying cell-based regenerative therapies is key in treating DKD. MSCs protect the kidneys from damage through multiple pathways involving autonomously targeted, anti-apoptotic, anti-inflammatory, antioxidant, and anti-fibrotic effects and podocyte autophagy regulation [44,45] The mechanism of this therapy is mainly achieved through two pathways: the paracrine action of stem cells and the exosomes secreted by stem cells [24,46]. First, MSCs reduce the expression of transforming growth factor β1 (TGFβ1) and inhibit the transdifferentiation of glomerular cells into myofibroblasts, which is a key pathological process in renal fibrosis. In addition, MSCs reduce the abnormal proliferation of glomerular cells by inhibiting the activation of phosphatidylinositol 3-kinase/Akt and mitogen-activated protein kinase signaling pathways, which are key factors in extracellular matrix (ECM) accumulation and glomerular expansion in DN. MSCs can also increase the expression of matrix metalloprotein 2 (MMP2) and MMP9, promote the degradation of ECM proteins, and reduce excessive accumulation of ECM. Simultaneously, MSCs secrete various cell growth factors, such as epidermal growth factor, which reduce the apoptosis of podocytes induced by hyperglycemia and promote the repair and regeneration of podocytes. Second, stem cells play a therapeutic role by secreting exosomes. Exosomes contain a variety of microRNAs (miRNAs) and mRNAs that regulate gene expression in target cells. For example, miR-21 inhibits the expression of programmed cell death protein 4 and reduces TGF-β-induced fibrosis. miR-192 and miR-215 downregulate E-cadherin expression and alleviate renal fibrosis. Exosomes transfer their contents to damaged tissues, promote the proliferation of glomerular and tubular epithelial cells, inhibit apoptosis, and repair damaged kidney tissues. Exosomes also inhibit the inflammatory response, reduce the infiltration of inflammatory cells and the production of inflammatory factors, and reduce the inflammation of the glomeruli and renal tubules [47–49].

The main challenges in applying SCT in patients with DKD are safety and efficacy. Although stem cell injections have a good overall safety profile for DKD, associated adverse events have been reported. The clinical studies in this review reported adverse events during SCT of DN, including asthma, atrioventricular block, fever, and diarrhea. During the study period, 36 events were reported in 46 patients (36/46, 78%) in the experimental group and 31 events were reported in the control group of 39 patients (31/39, 79%), all of which were of mild or moderate severity. Although the SCT and control groups showed a high incidence of adverse events, most were unrelated to SCT. This meta-analysis showed no significant difference in the incidence of adverse events between the SCT and the control groups (RR = 0.94,95% CI: 0.76–1.16; p = 0.56).

### 4.1 Limitations

The study had a few limitations. First, the number of RCTs included in this study was small, possibly contributing to the risk of not accounting for all findings. Second, most clinical studies of stem cells are still in their early stages, and stem cell isolation, purification methods, and injection routes vary, showing the lack of effective strategies to precisely target stem cells to damaged tissues in clinical trials. Different transplantation methods impact the survival and homing rate of MSCs, and the optimal implantation method, timing of treatment, and number of injections should be determined. Third, the results of the analysis included only the trial-level data. Only the main trial results were considered. Individual patient data were not available; this data could help determine whether the benefits of stem cells are limited to patients with DKD. Our subgroup analysis was based on eGFR, and its reliability will improve as individual patient data availability increase. Fourth, for the participants of the included trials, the progression from DKD to ESKD may take years to decades, a period that can vary widely between patients, affected by their baseline renal function, glycemic control, blood pressure management, and lifestyle habits. Trial participants were at different stages of DKD during the follow-up period; therefore, they may respond differently to cell therapy. This inevitably creates a bias in the results of the meta-analysis. Future studies should include more randomized controlled studies with large samples and individuals at the same or similar stage of injury to verify our conclusions.

## 5. Conclusion

The results of this study suggest that SCT can serve as a potential treatment modality for DKD and that it can significantly improve eGFR, decrease SCr, and reduce MAU, thus reducing renal damage. However, this study also showed that SCT was not effective in improving UACR levels. Owing to the obvious heterogeneity between the included studies, our results should be verified in RCTs with large sample sizes.

## Data Availability

All relevant data are within the manuscript and its Supporting Information files.

## Supplementary Materials

The following supporting information can be downloaded at: www.mdpi.com/xxx/s1, Figure S1: Forest plot for metabolic parameters; Table S1: Search Strategies for All Databases

## Author Contributions

Conceptualization, H-Y.D. and Y.Y.; methodology, H-Y.D.; software, C.X. ; validation, Z.L., Y.Y. and J.L.; formal analysis, H-Y.D and C.X. ; investigation, W.L.; resources, W.L.; data curation, J.Y.; writing—original draft preparation, H-Y.D; writing—review and editing, H-Y.D.; visualization, H-Y.D; supervision, H-Y.D and Z.L.; project administration, J.C.; funding acquisition, W.L. All authors have read and agreed to the published version of the manuscript.”

## Institutional Review Board Statement

Not applicable.

## Informed Consent Statement

Not applicable.

## Data Availability Statement

All authors agree to data sharing, and anyone who would like more data should contact the author at.

## Acknowledgments

The authors would like to thank the staff of Gannan Medical University for their valuable assistance.

## Disclaimer/Publisher’s Note

The statements, opinions and data contained in all publications are solely those of the individual author(s) and contributor(s) and not of MDPI and/or the editor(s). MDPI and/or the editor(s) disclaim responsibility for any injury to people or property resulting from any ideas, methods, instructions or products referred to in the content.

